# Adapting UK Biobank imaging for use in a routine memory clinic setting: the Oxford Brain Health Clinic

**DOI:** 10.1101/2022.08.31.22279212

**Authors:** Ludovica Griffanti, Grace Gillis, M. Clare O’Donoghue, Jasmine Blane, Pieter M. Pretorius, Robert Mitchell, Nicola Aikin, Karen Lindsay, Jon Campbell, Juliet Semple, Fidel Alfaro-Almagro, Stephen M. Smith, Karla L. Miller, Lola Martos, Vanessa Raymont, Clare E. Mackay

**Affiliations:** Department of Psychiatry, University of Oxford, UK; Oxford Health NHS Foundation Trust, Oxford, UK; Nuffield Department of Clinical Neurosciences, University of Oxford, UK; Wellcome Centre for Integrative Neuroimaging, University of Oxford, UK; Oxford University Hospitals NHS Trust, Oxford, UK

**Keywords:** UK Biobank, structural MRI, magnetic resonance imaging, memory clinic, dementia

## Abstract

The Oxford Brain Health Clinic (BHC) is a joint clinical-research service that provides memory clinic patients and clinicians access to high-quality assessments not routinely available, including brain MRI aligned with the UK Biobank imaging study (UKB).

In this work we present how we 1) adapted the UKB MRI acquisition protocol to be suitable for memory clinic patients, 2) modified the imaging analysis pipeline to extract measures that are in line with radiology reports and 3) compared measures from BHC patients to the largest brain MRI study in the world (ultimately 100,000 participants).

Adaptations of the UKB acquisition protocol for BHC patients include dividing the scan into core and optional sequences (i.e., additional imaging modalities) to improve patients’ tolerance for the MRI assessment. We adapted the UKB structural MRI analysis pipeline to take into account the characteristics of a memory clinic population (e.g., high amount of white matter hyperintensities and hippocampal atrophy). We then compared the imaging derived phenotypes (IDPs) extracted from the structural scans to visual ratings from radiology reports, non-imaging factors (age, cognition) and to reference distributions derived from UKB data.

Of the first 108 BHC attendees (August 2020-November 2021), 92.5% completed the clinical scans, 88.0% consented to use of data for research, and 48.1% completed at least one additional research sequence, demonstrating that the protocol is well tolerated. The high rates of consent to research makes this a unique real-world quality research dataset routinely captured in a clinical service. Modified tissue-type segmentation with lesion masking greatly improved grey matter volume estimation. CSF-masking marginally improved hippocampal segmentation. The IDPs were in line with radiology reports and showed significant associations with age and cognitive performance, in line with the literature. Due to the age difference between memory clinic patients of the BHC (age range 65-101 years, average 78.3 years) and UKB participants (44-82 years, average 64 years), additional scans on elderly healthy controls are needed to improve reference distributions. Current and future work aims to integrate automated quantitative measures in the radiology reports and evaluate their clinical utility.

## 1. Introduction

Brain magnetic resonance imaging (MRI) plays a key role in the diagnosis and evaluation of patients suspected of having dementia (Filippi et al., 2012; Staffaroni et al., 2017; Vernooij et al., 2019). Atrophy, specifically medial temporal lobe atrophy, has been included in the diagnostic guidelines for Alzheimer’s since 2011 (McKhann et al., 2011), and distinct atrophy patterns can point towards a specific underlying diagnosis of dementia subtype (Frisoni et al., 2010; McKhann et al., 2011; Staffaroni et al., 2017). More recently, there has been increasing awareness of the importance of vascular pathology as another key threat to cognitive health (Dickie et al., 2018; Gorelick et al., 2011; Smith et al., 2019).

Despite our understanding of structural changes in dementia, neuroimaging modalities are under-exploited in the clinical setting. In the UK, most people with memory problems (above 65 years old and with normal presentation) are referred to psychiatry-based memory clinics. These services typically do not have access to the same assessments (MRI and neuropsychology) as more specialist neurology-based clinics. Rather than to diagnose dementia, CT is most commonly used to exclude other pathologies, which account for a small proportion of cases (Scheltens et al., 2002; van Straaten et al., 2004). Specific atrophy patterns and the presence of vascular pathology are better assessed with MRI compared to CT (Scheltens et al., 2002; van Straaten et al., 2004).

The results of brain scans are then usually reported by a radiologist without the benefit of a standardised dementia-specific structure, leading to highly variable content across patients. Consortia and working groups are making an effort to standardise the structure of radiology reports, include information about vascular pathology, and often suggest the use of semi-quantitative scales (Filippi et al., 2012; Smith et al., 2019; Vernooij et al., 2019). Including quantitative measures extracted from MRI in radiology reports has the potential to increase the accuracy of dementia diagnosis and prognosis (Bosco et al., 2017; Vernooij et al., 2018), reduce inter-rater variability, and improve workflows (Goodkin et al., 2019). However, despite being widely used in dementia research, MRI and quantitative measures derived from MRI are not commonly used in routine clinical assessments (Vernooij et al., 2019).

In research settings, various neuroimaging software suites have been developed to derive quantitative measures from brain MRI (e.g. cortical and hippocampal volume, volume of white matter hyperintensities – WMHs) and have been widely implemented in research projects. More recently, with imaging now being more regularly included in large population cohorts, there are efforts to standardise these measurements and make them useable by non-imaging experts to maximise the impact of brain health information. A good example is the UK Biobank (UKB) study, which will ultimately include multi-organ (including brain) imaging for 100,000 participants. This can be combined with lifestyle, health and genetic data to produce predictive models for late life brain health (Littlejohns et al., 2020; Miller et al., 2016). The brain MRI UKB image processing pipeline (Alfaro-Almagro et al., 2018) automatically extracts thousands of measurements, so-called imaging derived phenotypes (IDPs). The pipeline uses tools largely from the FSL and FreeSurfer software libraries, which are used by thousands of research laboratories world-wide. However, these tools have primarily been validated only in a research context. The applicability of these tools and the UKB pipeline to an unselected clinical population remains unclear.

The Oxford Brain Health Clinic (BHC)(O’Donoghue et al., 2022a) aims to address this disconnect between the research and clinical domains, whilst providing an ideal clinical setting to validate MRI analysis tools. A joint clinical-research service that opened in August 2020, the BHC augments current NHS psychiatry-led memory clinic services to give patients and clinicians access to high-quality assessments not routinely available, including MRI instead of CT. MRI sequences used in the BHC are identical to, or compatible with, the UK Biobank imaging study (UKB) (Miller et al., 2016), enabling us to directly compare and integrate BHC results with the larger UKB imaging dataset. Our long-term aim is to use large, rich population datasets (along with all of the lifestyle, health and genetic data), like UKB, to produce predictive models that are anchored to real-world outcomes (BHC). However, when applying the UKB protocol and pipeline in a memory clinic setting, some questions arise: 1) is the UKB acquisition protocol well-tolerated by patients? 2) is the UKB pipeline robust and generalisable to a real-life memory clinic population? 3) can UKB data be used as a reference population for memory clinic patients?

In this work we describe how we have adapted the UKB protocol for use in the BHC, and evaluated the tolerability of the BHC protocol for memory clinic patients both in terms of compliance and image quality. We assess the performance of the UKB analysis pipeline and optimise the necessary automated segmentation tools for use with this patient group. Finally, we compare the characteristics of BHC patients with UKB participants and discuss challenges and opportunities of using UKB as normative distributions and implementing quantitative measures in BHC reports.

## 2. Methods

### 2.1. Patient population

Patients from Oxfordshire who have been referred to Oxford Health NHS Foundation Trust Memory Clinics are triaged by the duty Psychiatrist (RM) for referral to the Brain Health Clinic for assessments prior to their memory clinic appointment. Selection at the triage stage is based on clinical need and MRI safety screening. The GP referral and notes are examined for evidence of MRI safety contraindications and the duty psychiatrist speaks to the patient by phone to assess suitability for scanning. There is no explicit age cut-off for a BHC referral, but a clinical decision is made about the benefit of a BHC visit for patients who are deemed too frail for advanced scanning or where dementia is well established.

At the BHC, patients undergo extensive brain health assessments, including cognitive assessments, physiological measurements, questionnaires, and a 3T MRI scan.

Patients are also offered three ways to take part in research: 1) to share their clinical data for research use, 2) to undergo additional assessments during their visit and 3) to be informed about future opportunities to participant in studies and trials. All research is optional, and patients that choose not to take part in research still complete the NHS assessment at the BHC. The BHC Research Database was reviewed and approved by the South Central – Oxford C research ethics committee (SC/19/0404).

For more details on the non-imaging assessments performed and the research consent process, please refer to (O’Donoghue et al., 2022a).

### 2.2. BHC MRI protocol

The UKB brain MRI protocol was optimised to produce high-quality images in as short a time as possible, to accommodate the large N (Miller et al., 2016; Smith, 2020), making it ideal as a clinical protocol.

The BHC brain MRI protocol was designed to match as closely as possible the UKB, with some adaptations intended to make the scan more tolerable for memory clinic patients and to prioritise the collection of images that are currently most useful to aid dementia diagnosis. Table 1 illustrates the BHC MRI protocol compared with the UKB pipeline. The full protocol details are openly available on the WIN MR Protocols Database (O’Donoghue et al., 2022b).

**Table 1.**
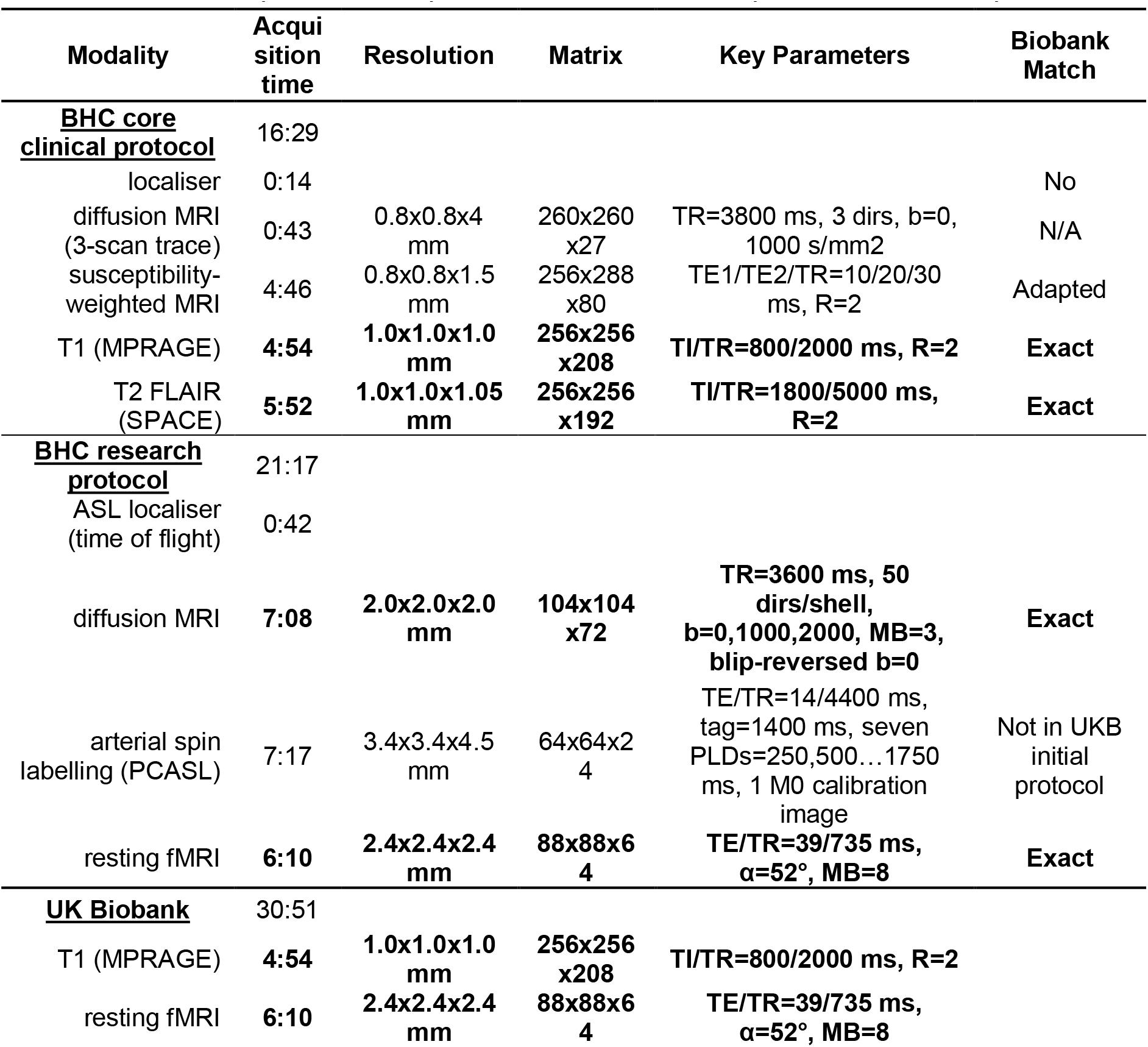
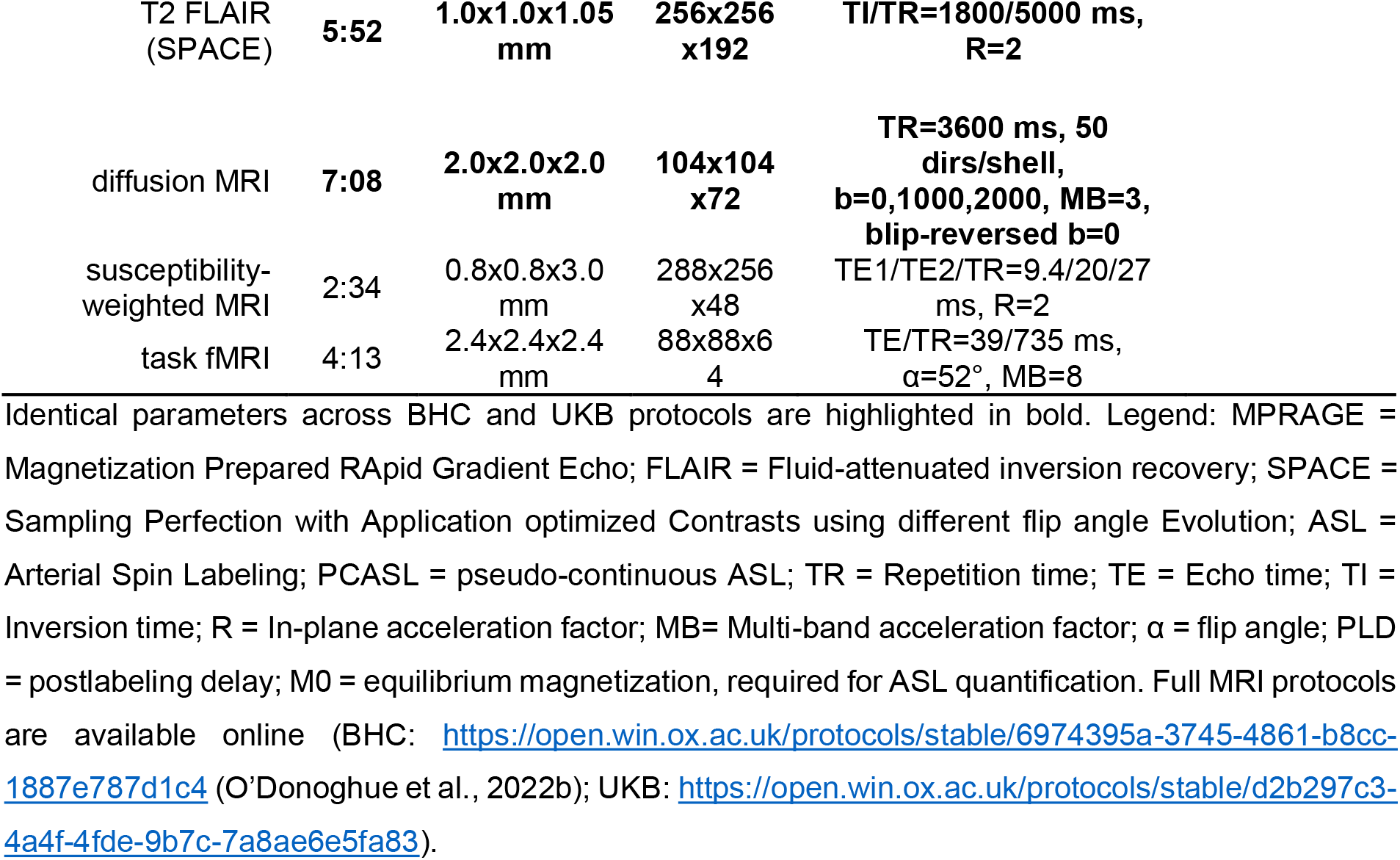
BHC MRI protocol: acquisition details and comparison with UKB protocol.

| Modality | Acquisition time | Resolution | Matrix | Key Parameters | Biobank Match |
| --- | --- | --- | --- | --- | --- |
| **<u>BHC core clinical protocol</u>** |  |  |  |  |  |
| localiser | 0:14 |  |  |  | No |
| diffusion MRI (3-scan trace) | 0:43 | 0.8x0.8x4 mm | 260x260 x27 | TR=3800 ms, 3 dirs, b=0, 1000 s/mm<sup>2</sup> | N/A |
| susceptibility-weighted MRI | 4:46 | 0.8x0.8x1.5 mm | 256x288 x80 | TE1/TE2/TR=10/20/30 ms, R=2 | Adapted |
| T1 (MPRAGE) | 4:54 | 1.0x1.0x1.0 mm | 256x256 x208 | TI/TR=800/2000 ms, R=2 | Exact |
| T2 FLAIR (SPACE) | 5:52 | 1.0x1.0x1.05 mm | 256x256 x192 | TI/TR=1800/5000 ms, R=2 | Exact |
| **<u>BHC research protocol</u>** |  |  |  |  |  |
| ASL localiser (time of flight) | 0:42 |  |  |  |  |
| diffusion MRI | 7:08 | 2.0x2.0x2.0 mm | 104x104 x72 | TR=3600 ms, 50 dirs/shell, b=0,1000,2000, MB=3, blip-reversed b=0 | Exact |
| arterial spin labelling (PCASL) | 7:17 | 3.4x3.4x4.5 mm | 64x64x24 | TE/TR=14/4400 ms, tag=1400 ms, seven PLDs=250,500...1750 ms, 1 M0 calibration image | Not in UKB initial protocol |
| resting fMRI | 6:10 | 2.4x2.4x2.4 mm | 88x88x64 | TE/TR=39/735 ms, $\alpha=52^\circ$, MB=8 | Exact |
| **<u>UK Biobank</u>** |  |  |  |  |  |
| T1 (MPRAGE) | 4:54 | 1.0x1.0x1.0 mm | 256x256 x208 | TI/TR=800/2000 ms, R=2 |  |
| resting fMRI | 6:10 | 2.4x2.4x2.4 mm | 88x88x64 | TE/TR=39/735 ms, $\alpha=52^\circ$, MB=8 | |

|  |  |  |  |  |
| --- | --- | --- | --- | --- |
| T2 FLAIR<br>(SPACE) | 5:52 | 1.0x1.0x1.05<br>mm | 256x256<br>x192 | TI/TR=1800/5000 ms,<br>R=2 |
| diffusion MRI | 7:08 | 2.0x2.0x2.0<br>mm | 104x104<br>x72 | TR=3600 ms, 50<br>dirs/shell,<br>**b=0,1000,2000, MB=3,**<br>**blip-reversed b=0** |
| susceptibility-weighted MRI | 2:34 | 0.8x0.8x3.0<br>mm | 288x256<br>x48 | TE1/TE2/TR=9.4/20/27<br>ms, R=2 |
| task fMRI | 4:13 | 2.4x2.4x2.4<br>mm | 88x88x6<br>4 | TE/TR=39/735 ms,<br>$\alpha=52^\circ$, MB=8 |

Identical parameters across BHC and UKB protocols are highlighted in bold. Legend: MPRAGE = Magnetization Prepared RAPid Gradient Echo; FLAIR = Fluid-attenuated inversion recovery; SPACE = Sampling Perfection with Application optimized Contrasts using different flip angle Evolution; ASL = Arterial Spin Labeling; PCASL = pseudo-continuous ASL; TR = Repetition time; TE = Echo time; TI = Inversion time; R = In-plane acceleration factor; MB= Multi-band acceleration factor; $\alpha$ = flip angle; PLD = postlabeling delay; M0 = equilibrium magnetization, required for ASL quantification. Full MRI protocols are available online (BHC: https://open.win.ox.ac.uk/protocols/stable/6974395a-3745-4861-b8cc-1887e787d1c4 (O'Donoghue et al., 2022b); UKB: https://open.win.ox.ac.uk/protocols/stable/d2b297c3-4a4f-4fde-9b7c-7a8ae6e5fa83).

The scanner used in UKB is a Siemens Skyra 3T running software platform VD13A, with a 32-channel RF receive head coil. The BHC protocol was setup on a Siemens Prisma 3T running VE11C, with a 32-channel head coil. The Prisma is a slightly newer, higher-specification scanner than the Skyra and is running a newer software platform, hence is able to run the UKB sequences and protocol.

#### 2.2.1. Subdivision into clinical and research sequences

Of the MRI modalities included in the UKB protocol, some are routinely used in clinical settings, while others are currently used mainly for research purposes.

Leveraging the joint clinical-research nature of the BHC we divided the UKB protocol into two sets of sequences: a core clinical protocol including the sequences that are compatible with current radiological examinations of memory clinic patients, and a research protocol that patients can opt-in to receive if they consent to additional research assessments.

#### 2.2.2. BHC core clinical protocol

The core clinical sequences include a T1-weighted scan, a T2-weighted Fluid Attenuated Inversion Recovery (FLAIR) scan, a diffusion-weighted (dMRI) scan and a susceptibility-weighted (swMRI) scan.

The T1 and T2-FLAIR scans are exactly matched with UKB protocol. These high-resolution structural scans (1 mm isotropic) allow clear depiction of brain anatomy, with high contrast between grey and white matter (T1) and highlight alterations to tissue compartments typically associated with pathology (T2-FLAIR).

We then acquire a short dMRI scan (43 seconds) with just 3 orthogonal diffusion directions, as commonly done in clinical practice to evaluate mean diffusivity and detect areas of restricted diffusion for the assessment of ischaemic injury or prion disease.

The susceptibility-weighted sequence was modified from the UKB protocol to obtain higher resolution images (1.5 mm slice thickness vs 3 mm in UKB protocol), useful for the assessment of venous vasculature, microbleeds or aspects of microstructure (e.g., iron, calcium and myelin).

The acquisition time for the core clinical sequences is 21:17 mins.

#### 2.2.3. BHC research protocol

The research sequences include the UKB dMRI and resting state functional MRI (rfMRI) sequences, and an arterial spin labelling (ASL) sequence.

The UKB dMRI sequence includes 100 diffusion-encoding directions across 2 b-shells (50x b=1000 s/mm^2^, 50x b=2000 s/mm^2^) with multiband acceleration factor of 3. With respect to the clinical 3-scan dMRI, this sequence enables finer measurement of the random motion of water molecules to infer information about WM microstructural properties and delineate the gross axonal organisation of the brain.

The UKB rfMRI sequence includes 490 timepoints (volumes) acquired with 2.4-mm isotropic spatial resolution and TR = 0.735 s, with multiband acceleration factor of 8. Resting-state functional MRI measures changes in blood oxygenation associated with intrinsic brain activity (i.e., in the absence of an explicit task or sensory stimulus). During resting-state scans, subjects are instructed to look at a crosshair, blink normally and try not to fall asleep.

We did not include the task fMRI sequence from UKB as it was not designed for this type of population. Instead, given the impact of vascular risk factors and vascular pathology on dementia (de la Torre, 2012; Iturria-Medina et al., 2016), it was deemed particularly important to look at vascular brain health in the BHC. We therefore included an Arterial Spin Labelling (ASL) sequence to look at brain perfusion, preceded by a time of flight (TOF) acquisition to localise neck vessels. We used a 2D-multi-slice EPI readout, with pseudo-continuous ASL tag duration of 1400 ms, seven post labelling delays (PLDs=250,500…1750 ms), seven label/control pairs at each PLD, and one calibration image. During these scans, subjects are also instructed to look at a crosshair, blink normally and try not to fall asleep. The original UKB protocol did not include ASL, however this modality has been recently collected on the UKB COVID-19 study participants.

The acquisition time for the optional research sequences is 16:29 mins.

#### 2.2.4. Raw scan quality control

We visually inspected the raw images to identify low quality scans to give an indication of tolerability of the protocol (e.g., motion artifacts that would be indicative of the ability of a memory clinic patient to lay still in the scanner), as well as to identify scans that might be informative about robustness in evaluating the analysis pipeline.

### 2.3. Radiology reports

All clinical scans are reported by an experienced radiologist (PP) in a standardised way, following the guidelines emerged from a survey of the European Society of Neuroradiology (ESNR)(Vernooij et al., 2019).

Table 2 describes the fields of the structured report, as well as the variables that were derived from it for the patients who consented to the use of clinical data for research.

**Table 2.** Standardised radiology report structure.

| Section | Description | Derived variables |
| --- | --- | --- |
| Atrophy pattern |  |  |
| Generalised atrophy | Comment of degree of global brain atrophy | Yes/no, asymmetric (yes/no), severity (mild/moderate/severe) |
| Medial Temporal lobe Atrophy (MTA) | Comment on regional brain atrophy, including MTA scale | Scheltens score left and right MTA |
| White matter hyperintensities (WMH) | State if present or not. Fazekas score for microangiopathic change. Is the distribution likely to be microangiopathic change or is there is suspicion of an alternative cause such as multiple sclerosis or CADASIL? | Fazekas score total WMH, periventricular (PWMH), deep (DWMH), alternative cause (yes/no) |
| Microhaemorrhages | State if present or not. If present, comment on likely aetiology based on number and distribution (CAA, hypertension, previous trauma) | Presence (yes/no), count, location |
| Previous infarction or intracranial haemorrhage | Presence of previous lobar haemorrhage | Presence (yes/no), comments |
|  | Presence of previous lobar or lacunar infarction |  |
| Restricted diffusion suggesting prion disease | Presence of areas of restricted diffusion suggesting prion disease. | Yes/no |
| Important negatives | Presence of mass, tumours, extra-axial collection, hydrocephalus. | Mass/tumours (yes/no), extra-axial collection (yes/no), hydrocephalus (yes/no) |
| Other incidental pathologies | Mention specifically any clinically relevant incidental pathology (e.g. developmental brain malformation, sinonasal pathology, skull base pathology, orbital pathology) | Details |

Legend: CADASIL=Cerebral autosomal dominant arteriopathy with subcortical infarcts and leukoencephalopathy; CAA=Cerebral Amyloid Angiopathy

### 2.4. BHC MRI analysis: adapting the UKB pipeline to memory clinic population

The second component of UKB imaging implemented in the BHC is the image analysis pipeline. The pipeline (Alfaro-Almagro et al., 2018) automatically process the images and extracts imaging derived phenotypes (IDPs), standardised quantitative measures that can be easily used and interpreted also by non-imaging experts. Because of the well-matched acquisition protocol, we were able to apply the pipeline, normally used only in research, to extract quantitative information from clinical scans of BHC patients.

In this work we focused on obtaining the most clinically useful IDPs for dementia diagnosis (i.e. those relevant to the sections included in the standardised radiology report), from amongst the IDPs currently generated by the pipeline (Alfaro-Almagro et al., 2018). We therefore analysed T1-weighted scans and T2-FLAIR scans and extracted measures of global atrophy (total GM volume), hippocampal atrophy (hippocampal volume) and white matter change (volume of periventricular and deep WMHs).

We then tested whether the UKB pipeline applied to a real-life memory clinic population was successful in extracting the measures of interest and, where necessary, performed the necessary adjustments to the analysis pipeline.

#### 2.4.2. Image processing with UKB pipeline

Structural images were analysed with FSL using the UKB analysis pipeline (version 1.5 - https://git.fmrib.ox.ac.uk/falmagro/UK_biobank_pipeline_v_1.5)(Alfaro-Almagro et al., 2018). In brief, the main steps of the T1 pipeline that derive the selected IDPs are: gradient distortion correction and defacing, brain extraction, linear and non-linear registration to standard space, FIRST subcortical structure segmentation (Patenaude et al., 2011), FAST bias field correction and tissue-type segmentation (Zhang et al., 2001) along with a shortened version of SIENAX reliant on FAST (Smith et al., 2002). The T2-FLAIR pipeline includes bias field correction, registration to T1 and standard space, brain extraction by masking with the T1 brain mask, and WMH segmentation with BIANCA (Griffanti et al., 2016), further subdivided into periventricular and deep WMH (Griffanti et al., 2018).

#### 2.4.3. UKB pipeline output quality check and optimisation

For the first 32 patients, visual inspection in FSLeyes was performed for the following stages in the UKB structural pipeline: brain extraction of T1 and T2-FLAIR scans, tissue-type segmentation from FAST/SIENAX (with particular attention to GM segmentation), hippocampus segmentation output from FIRST and WMH segmentation output from BIANCA. Each stage was rated as high, medium, or low quality by two raters independently (LG, GG) and the final rating was reached through consensus. Notes were included on the type of inaccuracy if present. These quality checks informed selection of the tools requiring optimisation.

Because this patient population is characterised by increased cortical and medial temporal lobe atrophy and presence of high white matter hyperintensities load, we identified two analysis steps that required optimisation (Figure 1).

**Figure 1.**
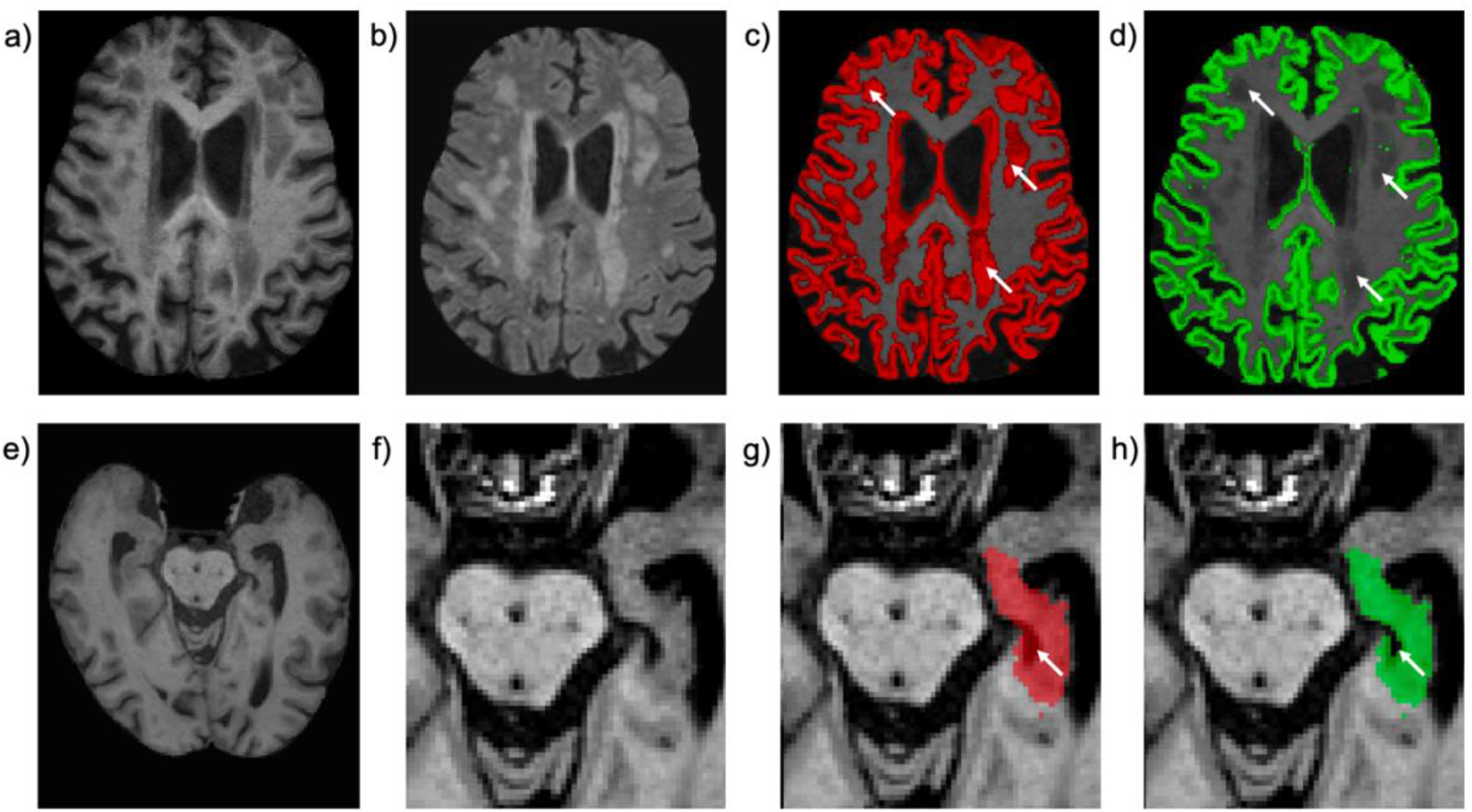
Results of UKB pipeline optimisation for memory clinic population. a) Example T1-weighted scan from a BHC patient where WMHs are T1 hypointense, similar in intensity to grey matter (GM). b) Corresponding T2-FLAIR scan showing WMHs. c) Uncorrected GM segmentation (red), with significant periventricular and deep WMHs classified as GM (white arrows). d) GM segmentation corrected with lesion-masking (green). e) Example T1-weighted scan from a different BHC patient, f) zoomed-in around the hippocampus. g) Uncorrected hippocampus segmentation (red) with errors (inclusion of CSF) highlighted by white arrows. h) CSF-masked hippocampus segmentation removed incorrectly labelled CSF voxels.

Grey matter (GM) segmentations (from FAST/SIENAX) were often flagged as inaccurate due to WMH being misclassified as GM, causing total GM volume to be overestimated. This is due to the fact that WMH that are bigger and likely at a more severe stage become visible on T1-weighted scans as hypo-intensities (Melazzini et al., 2021). To improve the segmentation accuracy, we implemented in the pipeline a modified version of SIENAX (-lm option). With this option WMHs (segmented with BIANCA) are initially excluded from tissue-type segmentation, then added to the final white matter map (since WMHs are, by definition, part of the WM). Visual inspection in FSLeyes was used to assess the accuracy of these strategies, and the most accurate segmentation strategy (original vs lesion-masking) was carried forward to subsequent analyses.

Hippocampal segmentations from FIRST were sometimes flagged as inaccurate in the quality checks. Since by design FIRST does not explicitly avoid the inclusion of non-grey tissues within its boundaries, one common error in this population with larger amounts of medial temporal lobe atrophy was the inclusion of cerebrospinal fluid (CSF) regions in the hippocampus. We therefore attempted CSF-masking to improve segmentation accuracy. In brief, this approach uses the CSF partial volume estimates (PVE) generated by FAST (thresholded to keep only voxels with high CSF content) to remove CSF areas from the FIRST hippocampal segmentations. The results of CSF-masking were visually inspected in FSLeyes. The most accurate segmentation strategy (with or without CSF masking) was carried forward in subsequent analyses.

#### 2.4.4. Optimised pipeline validation

We performed both an internal and external validation of the optimised analysis pipeline. Prior to any analyses, all IDPs were normalised for head size using the SIENAX scaling factor to correct for this between-subject variability. IDPs that were not normally distributed (Kolmogorov-Smirnov test) were cube-root transformed.

The internal validation was performed by examining the level of agreement between the IDPs extracted from the pipeline with visual ratings from radiology reports. We used ANOVA tests to compare the total GM volume across the 3-point scale of global cortical atrophy (none/mild/moderate or severe), the left and right hippocampal volumes obtained from FIRST with left and right medial temporal lobe atrophy (MTA) scale (Scheltens et al., 1992), and total WMH, PWMH and DWMH volumes from BIANCA against the Fazekas scale (Fazekas et al., 1987). If a visual rating score had fewer than 5 cases (usually lowest or highest scores), they were grouped with the nearest score for the statistical analyses.

External validation was then performed against non-imaging variables. Based on their known associations with WMHs and atrophy, three clinical variables were used for external validation of tool performance: patient age, the Addenbrooke’s Cognitive Examination-III (ACE-III)(Hsieh et al., 2013) total score and the ACE-III memory sub-score. Partial correlation (correlation with age controlled for sex, correlation with ACE-III and ACE-III memory score controlled for age and sex) was used to test the relationship between these variables and GM volumes, FIRST hippocampal volumes, and BIANCA WMH volumes, under the hypothesis that strong correlations would indicate high tool performance.

All statistical analyses were performed in SPSS 27 (IBM) and Bonferroni-corrected for multiple comparisons.

### 2.5. Towards quantitative radiology reports for the BHC

Finally, we explored the use of UKB data as a reference population to aid derivation of individual prediction on BHC patients. This would allow incorporating quantitative measures into radiology reports, delivering the enhanced brain information through a decision support tool that is interpretable and useful to clinicians and patients. To this aim, we compared the characteristics of BHC patients with those of the UKB participants, both in terms of demographics and hippocampal volume using previously published nomograms derived from 19,793 generally healthy UKB participants (Nobis et al., 2019).

## 3. Results

### 3.1. Sample characteristics

From the opening of the BHC in August 2020 to November 2021, 108 patients attended their BHC appointment and 101 consented for use of clinical data for research (age range 65-101 years, average 78.3 years, 50.5% female (O’Donoghue et al., 2022a)). As shown in Figure 2, 92.5% (N=100) completed the clinical scans. In this work we will report and analyse data from the 88.0% (N=95) of the patients who consented to their data to be used for research. Fifty-one patients (47.2%) additionally completed at least one research scan and forty-seven (43.5%) completed all research scans.

**Figure 2.**
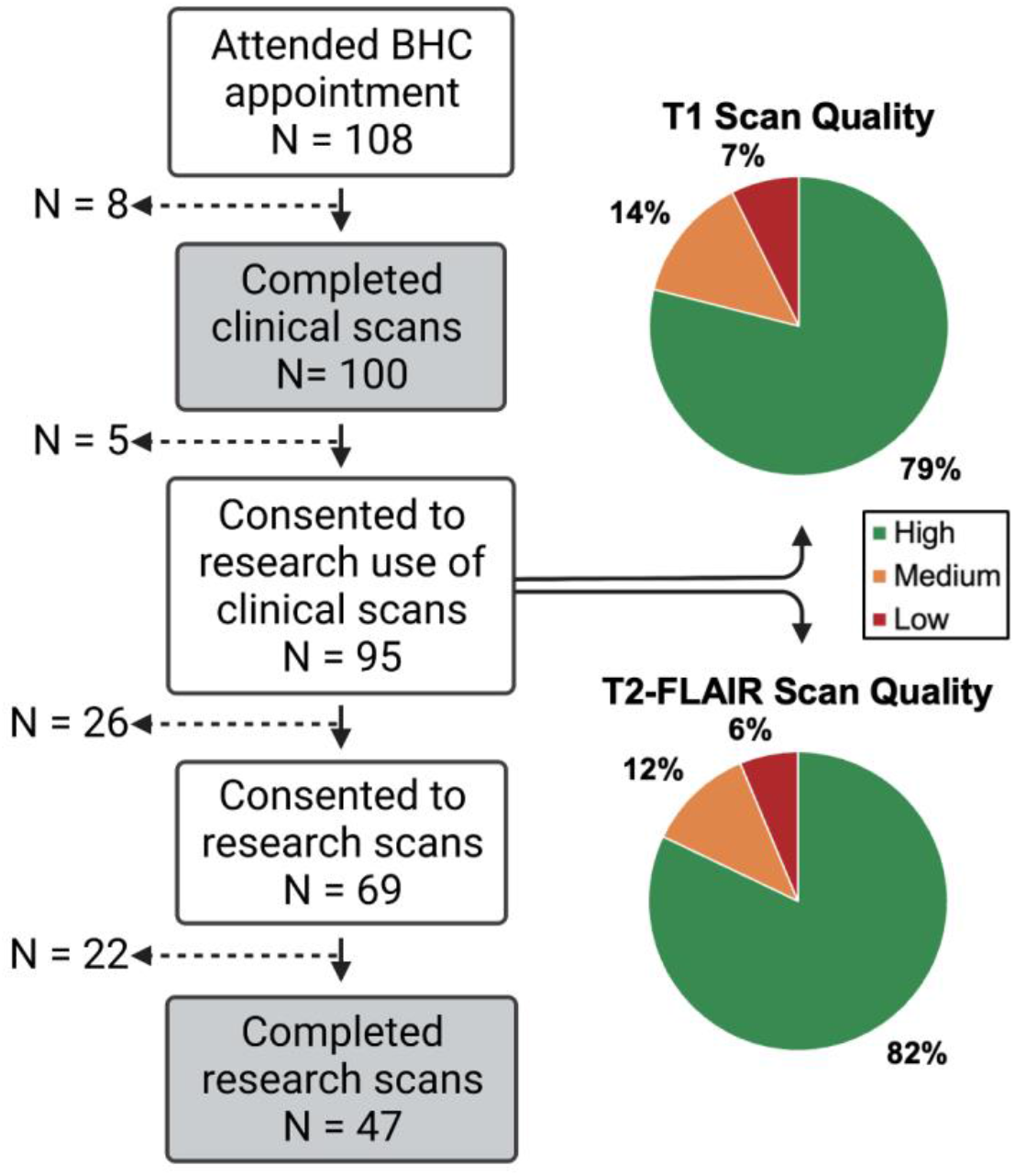
Flow chart of patients attending the Brain Health Clinic (consent and completion for clinical and research scans) and quality assessment of raw structural MRI data for the patients who completed the clinical scans and consented to the use of data for research (N=95).

Table 3 includes the characteristics of the 95 patients included, a summary of the findings on the radiology reports and the number of scans available for each modality.

**Table 3.** Sample characteristics

| **Demographics (N=95)** |  |
| --- | --- |
| Age (years) - mean ± SD (range) | 78.4 ± 6.2 (66-101) |
| Sex (M/F) | 47 / 48 |
| **Cognitive scores (N=92)** |  |
| ACE-III total score - mean ± SD (range) | 73.8 ± 17.1 (9-98) |
| ACE-III memory sub-score - mean ± SD (range) | 15.2 ± 6.5 (0-26) |
| **Radiology report (N=95)** |  |
| Generalised atrophy (counts for none/mild/moderate or severe) | 5 / 57 / 33 |
| L MTA score - median (range) [counts] | 2 (0-4) [6/26/32/27/4] |
| R MTA score - (range) [counts] | 2 (0-4) [6/28/29/29/3] |
| Fazekas PWMH score - median (range) [counts] | 2 (0-3) [3/44/33/15] |
| Fazekas DWMH score – median (range) [counts] | 1 (0-3) [4/54/36/1] |
| Fazekas total score – median (range) [counts] | 3 (0-6)<br>[1/5/38/17/19/14/1] |
| Microhaemorrhages (yes/no) | 10 / 85 |
| Previous haemorrhages (yes/no) | 3 / 93 |
| Previous infarcts (yes/no) | 8 / 88 |
| Restricted diffusion suggesting prion disease (yes/no) | 0 / 96 |
| Mass (yes/no) | 6 / 90 |
| Extra-axial collection (yes/no) | 0 / 96 |
| Hydrocephalus (yes/no) | 0 / 96 |
| **Clinical sequences (N=95)** |  |
| T1 | 95 |
| T2-FLAIR | 95 |
| dMRI 3-scan trace | 95 |
| swMRI | 95 |
| **Research sequences (N=51)** |  |
| dMRI | 51 |
| ASL | 51 |
| Resting fMRI | 47 |

Legend: SD= standard deviation; ACE-III = Addenbrooke's Cognitive Examination-III; MTA=Medial Temporal lobe Atrophy; PWMH=periventricular white matter hyperintensities; DWMH = deep white
matter hyperintensities; FLAIR=Fluid-Attenuated Inversion Recovery; swMRI=susceptibility-weighted imaging; dMRI=diffusion-weighted MRI; fMRI=functional MRI; ASL=Arterial Spin Labeling.

Quality checks of raw T1-weighted and T2-FLAIR images overall show that the UKB MRI sequences were suitable for this clinical population (93% of T1 scans and 94% of T2-FLAIR scans were high- or medium-quality, Figure 2). The most common quality issues were motion artifacts.

### 3.2. Radiology reports

A summary of the main findings from the radiology reports is provided in table 3.

We observed a level of generalised atrophy that was mostly mild or moderate. Out of the most severe cases, in two cases the atrophy level was reported as moderate to severe and in one case there was severe asymmetrical atrophy of the temporal lobe with relative sparing of the rest of the brain. The level of hippocampal atrophy was overall moderate (median MTA=2). The amount of white matter hyperintensities was predominantly moderate in the periventricular areas and mild in the deep white matter.

Three patients had a previous haemorrhage, and 8 patients had a previous infarct (one acute, seven chronic). Out of the ten patients presenting microhaemorrhages, in four cases there was a single lesion reported, in three cases there were two lesions, and in the remaining cases there where several lesions (more than five). A mass was reported for six patients: in 3 cases it was a cyst, 2 cases of meningioma, 1 cavernous haemangioma. No cases of restricted diffusion, extra-axial collection or hydrocephalus were reported.

### 3.3. BHC MRI analysis: pipeline optimisation and validation

The automated analysis pipeline failed on one participant (unsuccessful T1 brain extraction and registrations, preventing further analyses on other modalities), due to high levels of motion in the T1 scan (also marked as low-quality image in the raw data QC and mentioned in the radiology report as a low-quality scan). Results are therefore reported for the remaining 94 patients.

#### 3.2.1 UKB pipeline output quality check and optimisation

Table 4 shows the results of the visual check on the pipeline output on the first 32 patients before and after optimisation. The low number of high-quality tissue-type and hippocampal segmentations with the default pipeline prompted the need for optimising these processing steps. Although several WMH segmentations were rated of medium quality, the most common errors were small false positive clusters in the cortex and overestimation of the lesion size in cases with low WMH load. While there is still room for improvement in WMH segmentation, this was not set out as a priority, as the inaccuracies would not have a big impact on the total volume (and on the lesion masking procedure used to improve tissue-type segmentation). Similarly, the other steps of the pipeline (brain extraction and registration) were deemed of high or medium quality, with small inaccuracies unlikely to significantly affect the calculation of most IDPs.

**Table 4.** UKB pipeline output quality check before and after optimisation on the first 32 BHC patients.

| Pipeline output | Before optimisation |  |  | After optimisation |  |  |
| --- | --- | --- | --- | --- | --- | --- |
|  | High | Medium | Low | High | Medium | Low |
| T1 brain extraction (BET) | 25 | 6 | 1 | - | - | - |
| Tissue-type segmentation (FAST/SIENAX) | 2 | 10 | 20 | 27 | 5 | 0 |
| Hippocampus segmentation (FIRST) | 13 | 17 | 2 | 18 | 13 | 1 |
| T2-FLAIR brain extraction (BET) | 25 | 6 | 1 | - | - | - |
| White matter hyperintensities (BIANCA) | 17 | 12 | 3 | - | - | - |

After implementing the two optimisation strategies, we observed that the quality of the results improved in both cases; lesion masking significantly improved the quality of tissue-type segmentation, while CSF masking only led to a modest improvement of hippocampal segmentation (with a threshold of the CSF PVE map of 0.7 empirically chosen as the one giving the best results). In both cases, the modifications to the pipeline were deemed successful and volumes extracted with the optimised pipeline were used in further analyses.

#### 3.2.2 Optimised pipeline validation

Figure 3 shows the results of the internal validation of the optimised pipeline. Visual rating scores derived from the radiology reports for atrophy and white matter hyperintensities are compared with the corresponding IDPs obtained with the automated pipeline (all normalised for head size, WMH volumes additionally cube-root transformed before entering statistical analysis).

**Figure 3.**
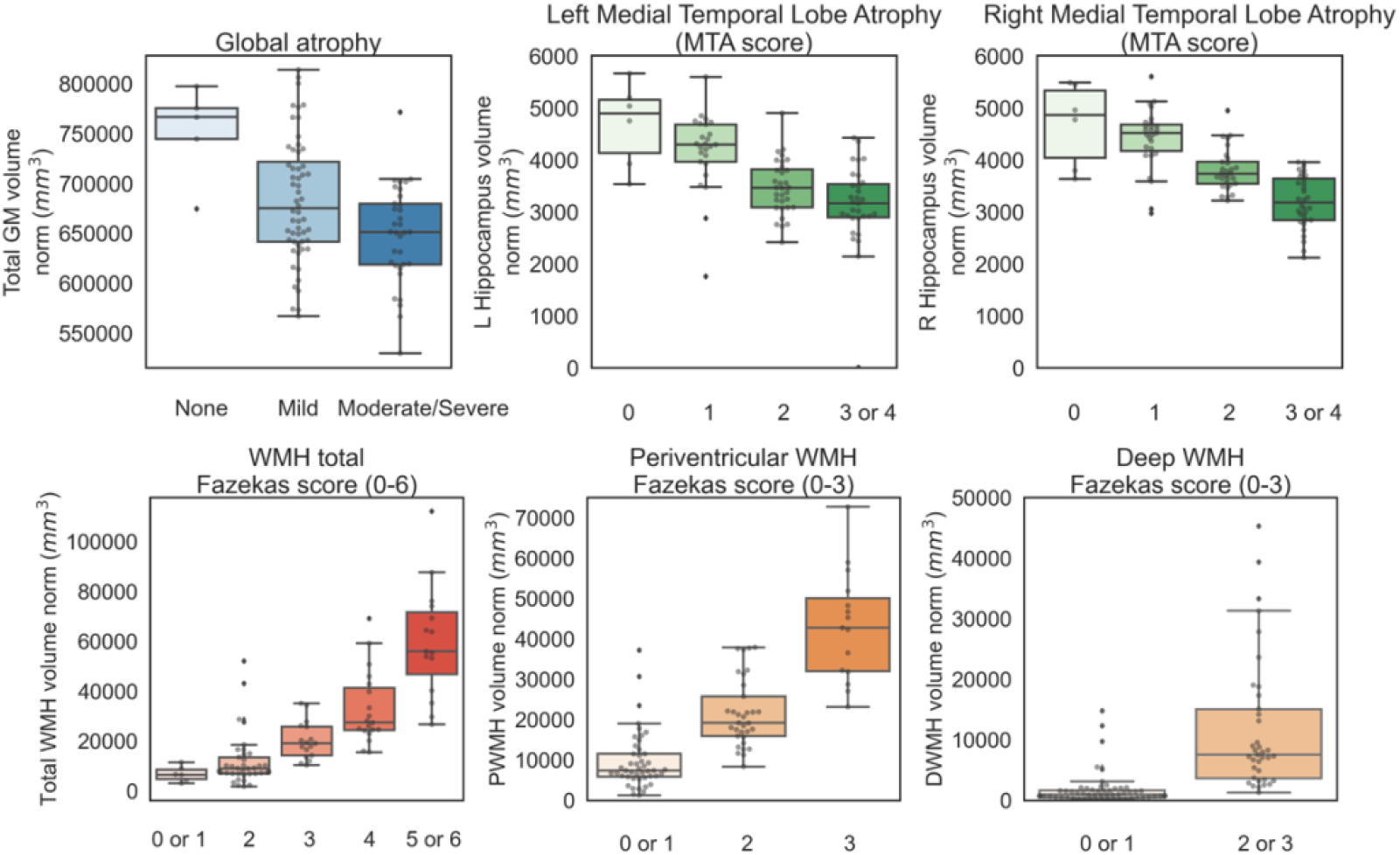
Optimised pipeline internal validation. Values of IDPs (y axes) derived from the automated pipeline, all normalised for head size (WMH volumes were cube-root transformed before entering statistical analyses) against the classifications (x axes) obtained from clinical radiology reports (blind to the results of the pipeline). Scores with less than 5 cases were grouped with the nearest score for statistical analyses (MTA 3 and 4; Total Fazekas 0 and 1, 5 and 6; PWMH Fazekas 0 and 1; DWMH Fazekas 0 and 1, 2 and 3). See table 3 for original count for each score and supplementary Figure 1 for boxplots with original scores).

The ANOVA tests showed that there was a statistically significant difference in total GM volume between the different atrophy categories (F(2,91) = 8.682, p < 0.001, partial eta squared = 0.160), a significant difference in hippocampal volumes across MTA scores (Left: F(3,90)=15.360, p<0.001, partial eta squared = 0.339; Right: F(3,90)=32.024, p<0.001, partial eta squared = 0.516) and a significant difference in white matter hyperintensity volumes across Fazekas scores (Total: F(4,89)=43.642, p<0.001, partial eta squared = 0.662; PWMH: F(2,91)=75.355, p<0.001, partial eta squared = 0.624; DWMH: H(1,92)=93.888, p<0.001, partial eta squared = 0.505). All results survive Bonferroni correction.

Table 5 reports the correlations between the IDPs and age (controlling for sex), total ACE III score and ACE III memory subscore (controlling for age and sex), performed as an external validation of the optimised pipeline.

**Table 5.**
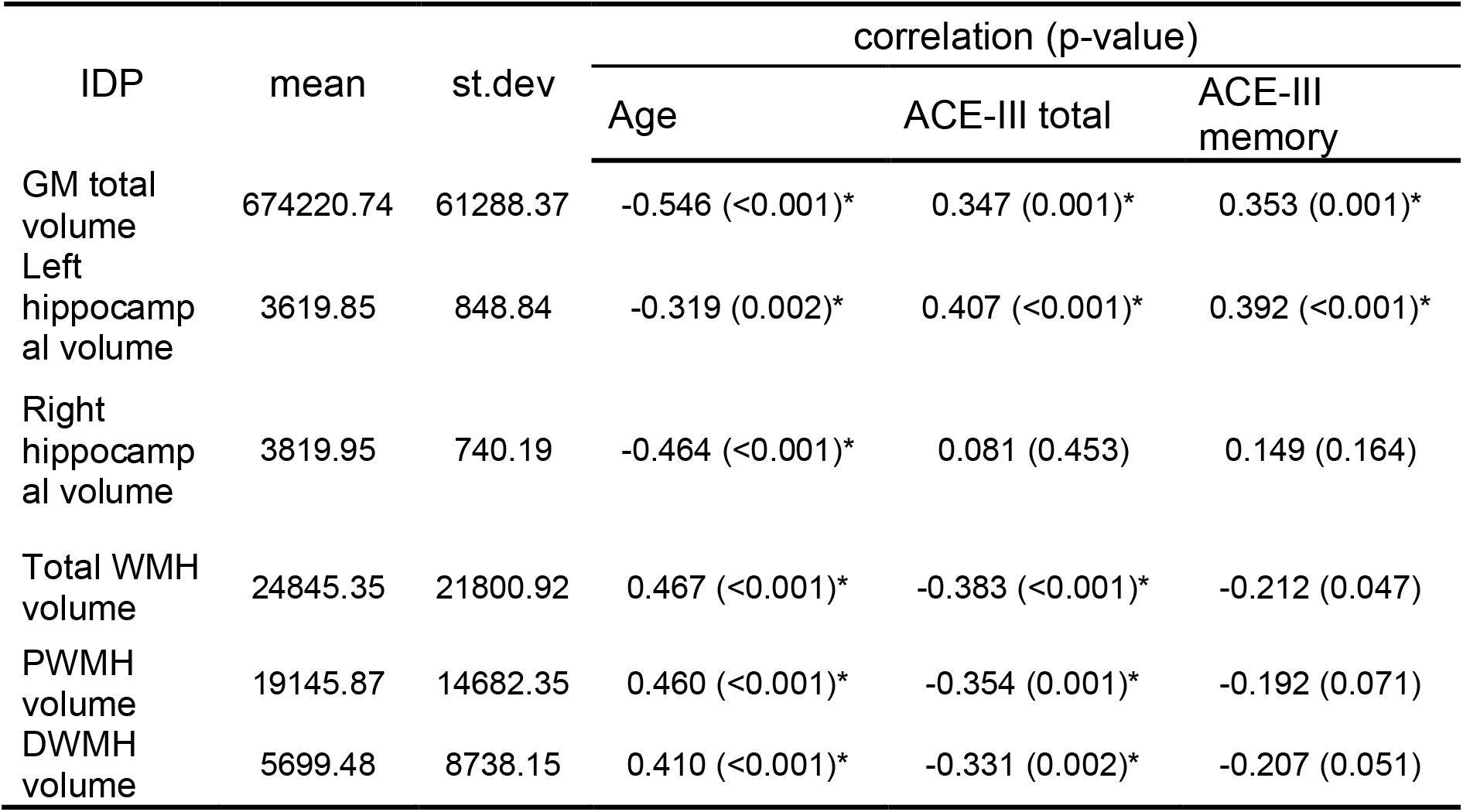
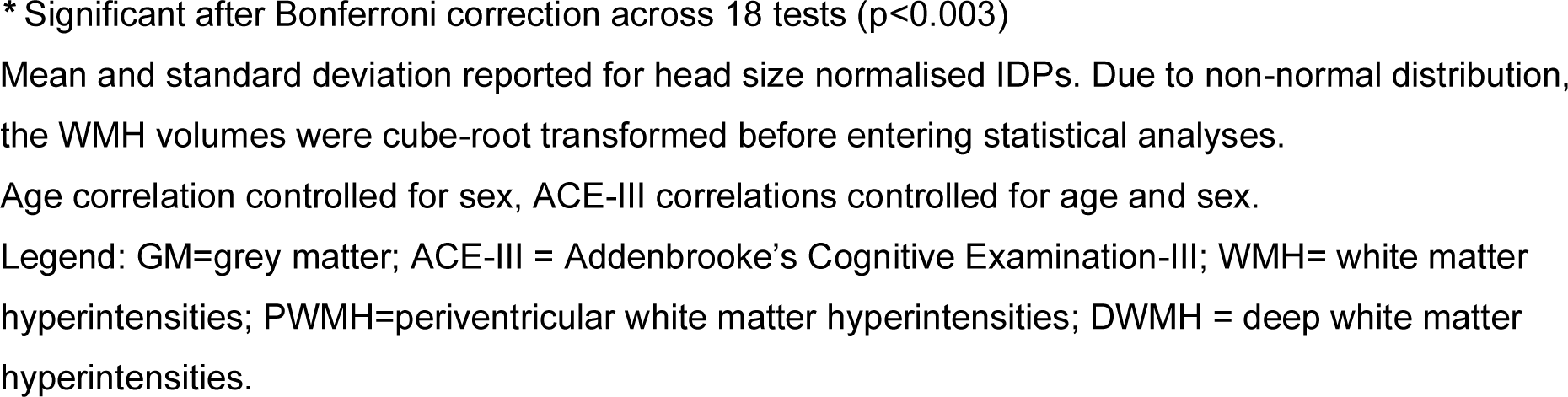
Optimised pipeline external validation.

| IDP | mean | st.dev | correlation (p-value) |  |  |
| --- | --- | --- | --- | --- | --- |
|  |  |  | Age | ACE-III total | ACE-III memory |
| GM total volume | 674220.74 | 61288.37 | -0.546 (<0.001)* | 0.347 (0.001)* | 0.353 (0.001)* |
| Left hippocampal volume | 3619.85 | 848.84 | -0.319 (0.002)* | 0.407 (<0.001)* | 0.392 (<0.001)* |
| Right hippocampal volume | 3819.95 | 740.19 | -0.464 (<0.001)* | 0.081 (0.453) | 0.149 (0.164) |
| Total WMH volume | 24845.35 | 21800.92 | 0.467 (<0.001)* | -0.383 (<0.001)* | -0.212 (0.047) |
| PWMH volume | 19145.87 | 14682.35 | 0.460 (<0.001)* | -0.354 (0.001)* | -0.192 (0.071) |
| DWMH volume | 5699.48 | 8738.15 | 0.410 (<0.001)* | -0.331 (0.002)* | -0.207 (0.051) |

\* Significant after Bonferroni correction across 18 tests ( $p < 0.003$ )
Mean and standard deviation reported for head size normalised IDPs. Due to non-normal distribution, the WMH volumes were cube-root transformed before entering statistical analyses.
Age correlation controlled for sex, ACE-III correlations controlled for age and sex.
Legend: GM=grey matter; ACE-III = Addenbrooke's Cognitive Examination-III; WMH= white matter hyperintensities; PWMH=periventricular white matter hyperintensities; DWMH = deep white matter hyperintensities.

Total GM volume showed significant negative correlation with age and positive correlation with ACE total and memory subscore. Regarding hippocampal volumes, significant negative correlations were observed between age and both the left and right hippocampi. Left hippocampal volume was also significantly correlated with ACE-III scores (total and memory sub-score), while correlations between right hippocampus and ACE-III scores were not significant. We further verified that the association between hippocampal volumes and ACE-III score was significantly different between hemispheres (F(1,89)=6.062, p=0.015 on interaction between hemisphere and ACE-III score using a linear mixed model - lmer package in R).

Greater WMH total, periventricular and deep volumes significantly correlated with higher age and lower ACE-III total score. None of the correlations between WMH volumes and ACE-III memory score reached Bonferroni-corrected significance.

### 3.4. Towards quantitative radiology reports for the BHC

Figure 4 shows how the BHC patients compare to 19,793 generally healthy UKB participants (Nobis et al., 2019). The age distribution of BHC and UKB participants is significantly different (BHC: 78.4 ± 6.2 years – range 66-101); UKB: 63.16 ± 7.50 years) years -range 45-81. Consequently, only a very small minority fall within the nomograms age range. For these patients, hippocampal volumes are overall comparable to those from UKB participants, with several patients falling in the lower percentiles.

**Figure 4.**
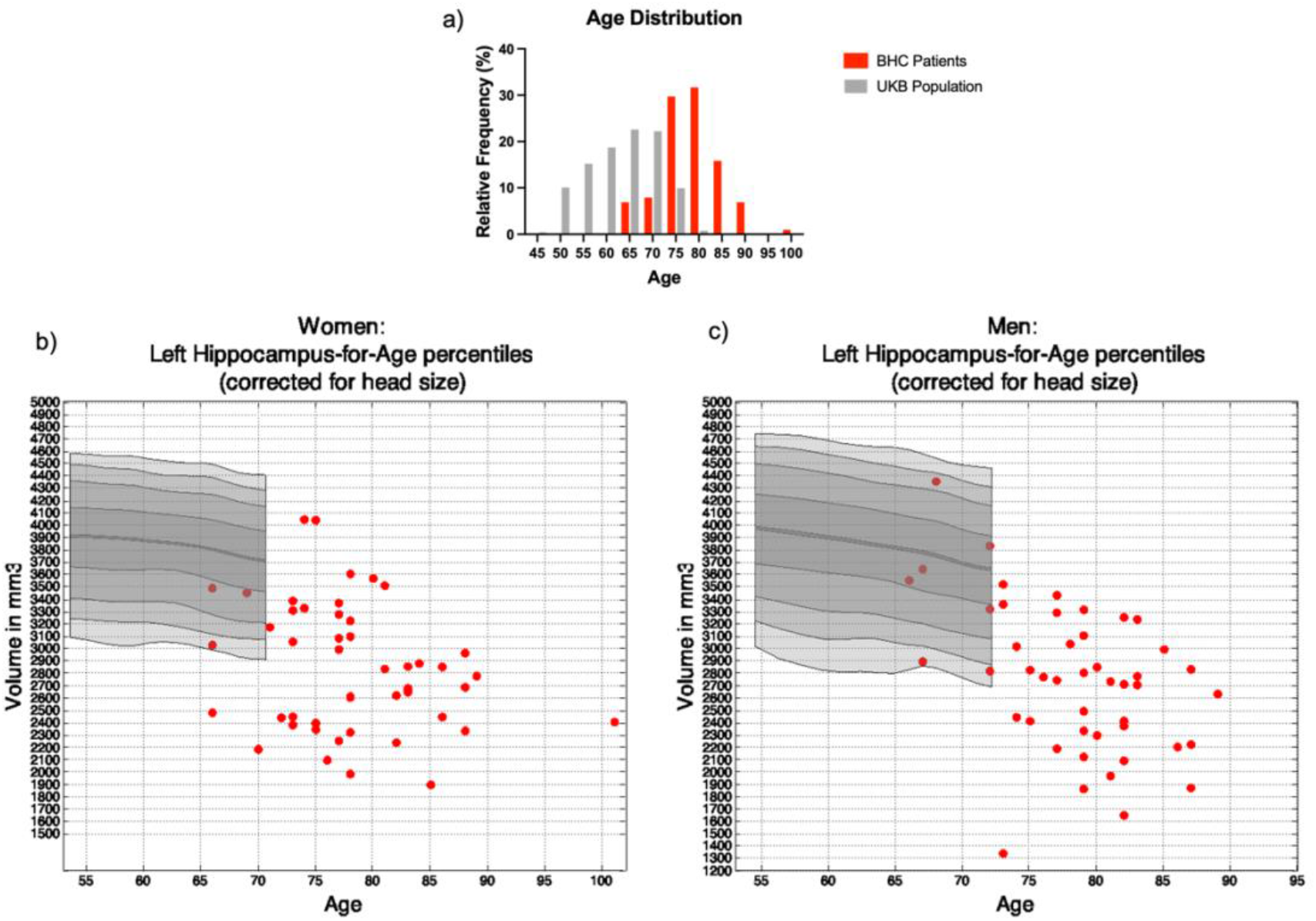
Comparison of BHC and UKB data. a) Histogram of age distribution for 94 BHC patients (red) and 19,793 UKB participants (grey). b) Left hippocampal volumes of female BHC patients (red) compared with UKB data (grey); c) Left hippocampal volumes of male BHC patients (red) compared with UKB data (grey nomogram). Most BHC patients fall in the lower percentiles of the nomograms, suggesting potentially pathological deviations from the norm. The remaining BHC patients were outside the nomograms age range, highlighting need for expanded normative data. See supplementary Figure 2 for right hippocampus results.

## 4. Discussion

In this work we described how we adapted the UKB brain imaging for use in a real-world memory clinic setting, the Oxford Brain Health Clinic.

The BHC acquisition protocol was found to be well-tolerated by patients. By dividing the UKB brain MRI protocol into clinical and research sections we were able to exploit its technical advances to generate high quality data in a reduced time, while tailoring it to inform dementia diagnosis. The high overall quality of raw T1-weighted and T2-FLAIR scans suggests that these UKB-matched sequences are suitable for patients with memory problems.

By combining clinical scans and assessments with optional research add-ons into a single session and enabling patients to opt for their desired level of research participation, this protocol reduces potential barriers to research engagement. Of those who were able to complete the clinical scans, 88% consented to the research use of their data. By prioritising data collection with direct clinical benefit and enabling additional research participation, this approach exceeds the target outlined in the UK Prime Minister’s Challenge on Dementia for 10% of dementia patients to participate in research (UK Department of Health, 2012). This results in a dataset that is representative of the memory clinic population, as the patients are not selectively recruited for a research study. Because patients can also consent to be recontacted for research (current consent rate 73.1% (O’Donoghue et al., 2022a)), this protocol is also building a valuable participant database for future studies.

The UKB analysis pipeline enables high-throughput automated generation of IDPs. However, because this pipeline was designed for and validated in a healthier population, the BHC group presents novel age- and disease-related challenges, including more hippocampal atrophy and WMHs.

In this unselected clinical population, tissue-type segmentation was highly affected by the presence of WMHs, misclassified as GM, due to their similar T1 intensity (Dadar et al., 2018; Melazzini et al., 2021). This inaccuracy has been reported elsewhere with multiple sclerosis (MS) lesions (Battaglini et al., 2012) and WMHs (Dadar et al., 2021). Lesion-filling (i.e. replacing or “filling” the intensity values in the lesion area with intensities that are similar to those in the non-lesion neighbourhood) and lesion-masking (the approach used in this study) are valuable correction tools for MS lesions (Battaglini et al., 2012; Chard et al., 2010). For WMHs of presumed vascular origin, we opted for the use of lesion-masking since WMHs tend to be larger and more confluent than the MS lesions for which lesion-filling was designed (Battaglini et al., 2012). Although alternative methods have been previously described to adjust for WMHs in GM segmentations (Park et al., 2018), lesion-masking provides a straightforward method yielding remarkable improvements in GM segmentation accuracy. Whilst essential in BHC patients with high WMH burdens, lesion-masking was not detrimental in patients with mild WMHs. Overall, the efficacy and simplicity of lesion-masking supports its widespread application in other pipelines and for other populations.

Regarding hippocampal segmentation, FIRST tended to either underestimate or, more commonly, overestimate hippocampal volume, particularly in patients with severe hippocampal atrophy. This is consistent with the better performance of FIRST in non-atrophic brains (Goubran et al., 2020), despite the inclusion of some AD patients in its training datasets (Patenaude et al., 2011). Similar inaccuracies have been reported with other hippocampal segmentation tools (Firbank et al., 2008), supporting the widespread need for optimisation. CSF-masking, a novel strategy in this study, led to a modest improvement of the segmentations, despite very small impact on the calculated volumes. This may be because some segmentation inaccuracies also occur at the GM-WM interface, and future efforts should aim to also improve segmentation accuracy at this boundary. Another possibility is that FIRST segmentations may be inaccurate in shape or location but not necessarily in size. In fact, among the segmentations that were still labelled as low quality after CSF-masking, only one was an outlier in terms of volume. Nevertheless, further optimisation is required to improve the quality of these segmentations, especially for future analyses reliant on hippocampal shape (e.g., vertex analysis).

Unlike many other studies that evaluate tool performance in selected patient groups or healthy controls, here we validated automated segmentation tools in an unselected clinical population. Because every tool validation method has limitations, we used both internal and external validation strategies.

For our internal validation (i.e., using imaging information) we compared IDPs of total GM volume, hippocampal volume and WMH (total, periventricular and deep) with the corresponding scales from the radiology reports. We found very good agreement for all IDPs, suggesting that the pipeline can be used to automatically extract meaningful information from memory clinic patients.

For the external validation (i.e., using non-imaging information) we correlated the same IDPs with clinical factors known to be associated with atrophy and WMHs: age, and cognition (using ACE-III total score and memory subscore). While age was significantly correlated with reduced GM and hippocampal volume as well as higher WMH volumes, ACE-III score was significantly associated with GM volume, left hippocampal volume, and WMH volumes. The memory subscore of ACE-III was only associated with GM and left hippocampal volume, but not WMH volumes.

The significant difference in correlations between the left and right hippocampus could be due to the fact that the volume of the left hippocampus has been previously found to be smaller and more strongly correlated with cognitive symptoms in MCI and dementia than the right hippocampus (Ezzati et al., 2016; Muller et al., 2005; Shi et al., 2009). Compared to the right hippocampus, we see smaller left hippocampal volumes with greater variability both before and after correction with CSF-masking (Table 5, paired t-test L<R hippocampus p=0.003). This smaller variability may also limit our power to detect correlations with ACE-III total and memory scores on the right side.

Also regarding WMH volumes our results are in agreement with previous studies in older adults, which found PWMH to be associated with impaired cognitive function (Bolandzadeh et al., 2012; Griffanti et al., 2018; Kim et al., 2008), although we also found significant association in DWMH.

Finally, thanks to the use of UKB acquisition protocol and pipeline, we could compare BHC patients with UKB participants, to explore the use of UKB data as reference for individual predictions in memory clinic patients. In particular, we used previously published nomograms for hippocampal volume (Nobis et al., 2019) to assess where individual BHC patients fall in respect to a wider population of the same age and sex. The results showed that the substantial age difference between UKB participants and BHC patients make UKB data currently not suitable to be used as reference population for memory clinic patients. Nomograms used here were generated using 19,793 participants, although data from the latest release including 51,532 participants who underwent imaging (visit 2 – see supplementary figure 3) are very similar, with a mean age of 64.54 ± 7.81 and age range between 44 and 83 years. The use of a sliding window approach further restricts the age range, since the nomograms were designed so that each window contained 10% of the participants (Nobis et al., 2019). Using alternative fitting methods while considering the increase in uncertainty when fewer samples are available (e.g. either side of the age range), may help building nomograms that cover a wider age range.

More recently, brain charts of brain tissues were generated from over 100,000 scans (including UKB) across the lifespan (Bethlehem et al., 2022). They include the age range of BHC patients, but being generated from multi-site data, they currently require at least 100 scans from healthy controls to estimate study-specific offset. Moreover, these charts are not yet extended to more fine-grained IDPs that are likely to be more useful for dementia diagnosis, but also more sensitive in variations in image quality.

To overcome these limitations, we have now started to acquire additional scans on elderly healthy controls with the same protocol to improve reference distributions and to be able to detect pathological deviations.

Additional future work includes expanding the UKB pipeline to detect other brain characteristics that are currently assessed in the clinical setting and that would be useful to quantify automatically (e.g., other signs of small vessel disease like lacunes on FLAIR scans or microhaemorrhages on swMRI scans).

On the other hand, the UKB pipeline (including all the sequences) currently produces thousands of IDPs, most of which are not related to brain characteristics currently assessed in clinical settings. Having access to such data from a representative population will allow us to test their clinical utility individually and in combination with non-imaging variables (e.g. lifestyle factors, health measures and genetics).

Ultimately the IDPs and relative reference distributions could be integrated to produce an imaging decision support tool for dementia diagnosis, the clinical utility of which can be directly assessed in the clinical setting at the BHC.

To conclude, to the best of our knowledge, this is the first time that UKB imaging (acquisition, analysis pipeline and reference data) is used on an unselected real-world patient population. This demonstrates the possibility to bridge the gap between research and clinical practice and lead the way for integrated research and clinical assessments.

## Data Availability

The MRI data presented in this paper will be available via the Dementias Platform UK (https://portal.dementiasplatform.uk/CohortDirectory/Item?fingerPrintID=BHC) and access will be granted through an application process, reviewed by the BHC Data Access Group. The BHC Data Access Group will start accepting applications to access BHC data upon peer-reviewed publication of the present work. Data will continue to be released in batches as the BHC progresses in order to minimise the risk of participant identification.
The complete BHC MRI protocol and scanning procedure is available through the WIN MR Protocols Database at: https://open.win.ox.ac.uk/protocols/stable/6974395a-3745-4861-b8cc-1887e787d1c4.
The UK Biobank brain MRI analysis pipeline used in this study (v1.5) is openly available (https://git.fmrib.ox.ac.uk/falmagro/uk_biobank_pipeline_v_1.5/-/tree/master). Modified or additional scripts for the analyses performed in this study are available at (https://git.fmrib.ox.ac.uk/open-science/analysis/brain-health-clinic-mri)

## Data/Code availability statement

The complete BHC MRI protocol and scanning procedure is available through the WIN MR Protocols Database at: https://open.win.ox.ac.uk/protocols/stable/6974395a-3745-4861-b8cc-1887e787d1c4 (O’Donoghue et al., 2022b).

The UK Biobank brain MRI analysis pipeline used in this study (v1.5) is openly available (https://git.fmrib.ox.ac.uk/falmagro/uk_biobank_pipeline_v_1.5/-/tree/master). Modified or additional scripts for the analyses performed in this study are available at (https://git.fmrib.ox.ac.uk/open-science/analysis/brain-health-clinic-mri)

The MRI data presented in this paper will be available via the Dementias Platform UK (https://portal.dementiasplatform.uk/CohortDirectory/Item?fingerPrintID=BHC) and access will be granted through an application process, reviewed by the BHC Data Access Group. The BHC Data Access Group will start accepting applications to access BHC data upon publication of the present work. Data will continue to be released in batches as the BHC progresses in order to minimise the risk of participant identification.

## Acknowledgements

Amanda Colston, Clare Hamblin, Emily Johnson, Rebecca Williams, Nicky Watkins, Jessica Wallis, Sophie Walker, Leona Wolters, Michael Ben-Yehuda, Emma Craig, Gary Gibbs, Karla Westphal, Rebecca Smith, Deborah Wilkinson, Gemma Butler, Aurelija Burbaite, Julia Hamer Hunt, Qi Pei, Becci Dow, Candy Stone, Sebastian Rieger, Thomas Okell, Mark Chiew, Paul Semple, Shona Forster, Thomas Nichols.

## CRediT author statement

*Ludovica Griffanti* – Conceptualization; Methodology; Software; Investigation; Data Curation; Formal analysis; Validation; Visualization, Writing - Original Draft

*Grace Gillis* – Methodology; Software; Investigation; Data Curation; Formal analysis; Validation; Visualization; Writing - Original Draft

*M. Clare O’Donoghue* - Conceptualization; Methodology; Investigation; Data curation; Project Administration; Writing – review & editing

*Jasmine Blane* - Investigation; Data curation; Project Administration, Writing – review & editing

*Pieter M. Pretorius* – Conceptualization, Investigation, Writing – review & editing

*Robert Mitchell* – Investigation, Writing – review & editing

*Nicola Aikin* - Investigation; Resources; Writing – review & editing

*Karen Lindsay* - Data curation; Project Administration, Writing – review & editing

*Jon Campbell* - Investigation; Resources; Writing – review & editing

*Juliet Semple* - Investigation; Resources; Writing – review & editing

*Fidel Alfaro-Almagro* - Methodology; Software; Writing – review & editing

*Stephen M. Smith* - Methodology; Software; Funding acquisition; Writing – review & editing

*Karla L. Miller* - Methodology; Software; Funding acquisition; Writing – review & editing

*Lola Martos* - Conceptualization; Resources; Funding acquisition; Writing – review & editing

*Vanessa Raymont* - Conceptualization; Resources; Funding acquisition; Supervision; Writing – review & editing

*Clare E. Mackay* - Conceptualization; Methodology; Resources; Funding acquisition; Supervision; Writing – review & editing

