## supplementary for "Adapting UK Biobank imaging for use in a routine memory clinic setting: the Oxford Brain Health Clinic"

### Supplementary figures

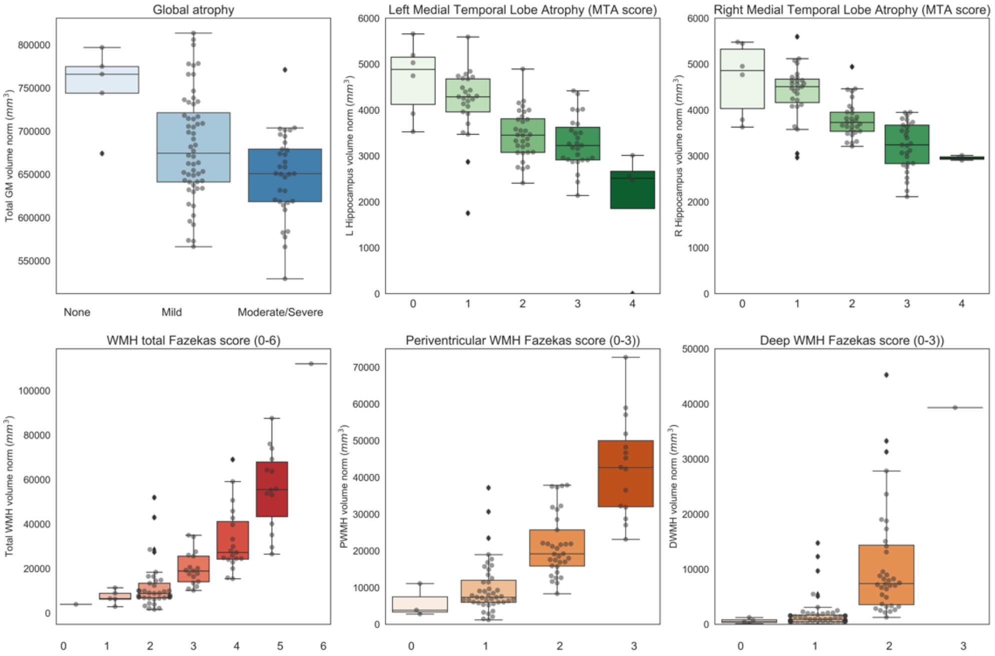


**Figure 1**. Optimised pipeline internal validation. Values of IDPs (y axes) derived from the automated pipeline, all normalised for head size against the classifications (x axes) obtained from clinical radiology reports (blind to the results of the pipeline). Before entering statistical analyses, scores with less than 5 cases were grouped with the nearest score, and WMH volumes were cube-root transformed, being not normally distributed (see main text).

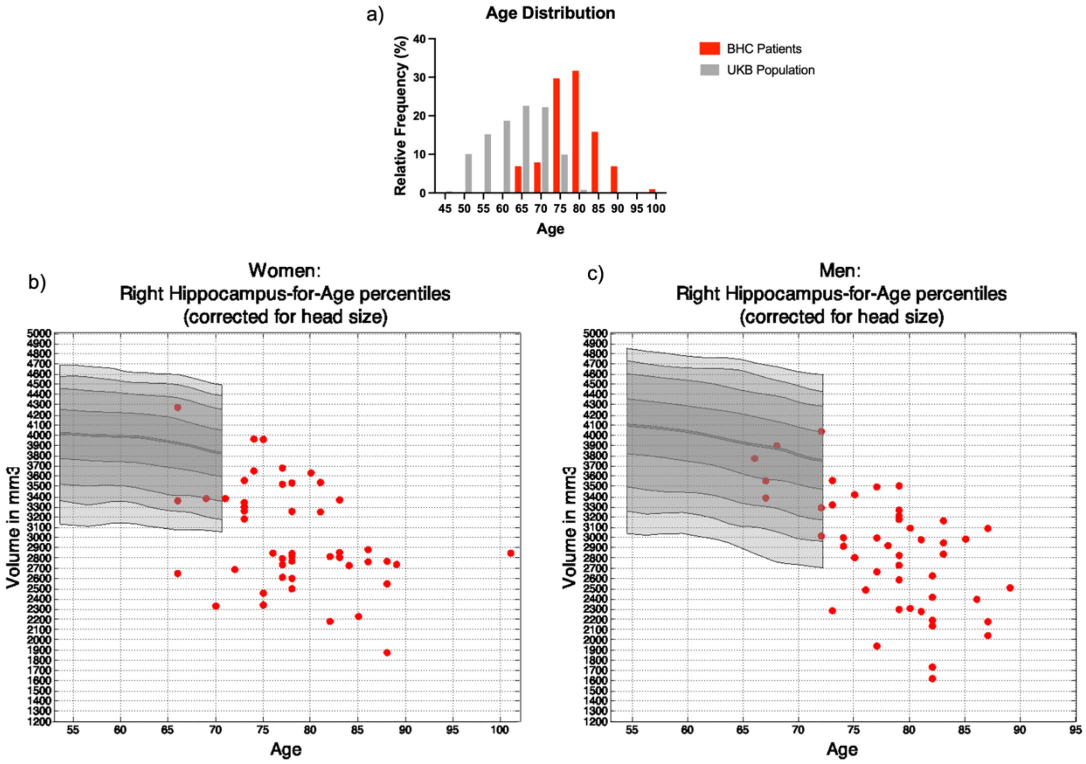


**Figure 2**. Comparison of BHC and UKB data. a) Histogram of age distribution for 94 BHC patients (red) and 19,793 UKB participants (grey). b) Right hippocampal volumes of female BHC patients (red) compared with UKB data (grey); c) Right hippocampal volumes of male BHC patients (red) compared with UKB data (grey nomogram). Most BHC patients fall in the lower percentiles of the nomograms, suggesting potentially pathological deviations from the norm. The remaining BHC patients were outside the nomograms age range, highlighting need for expanded normative data. See main text for left hippocampus results.

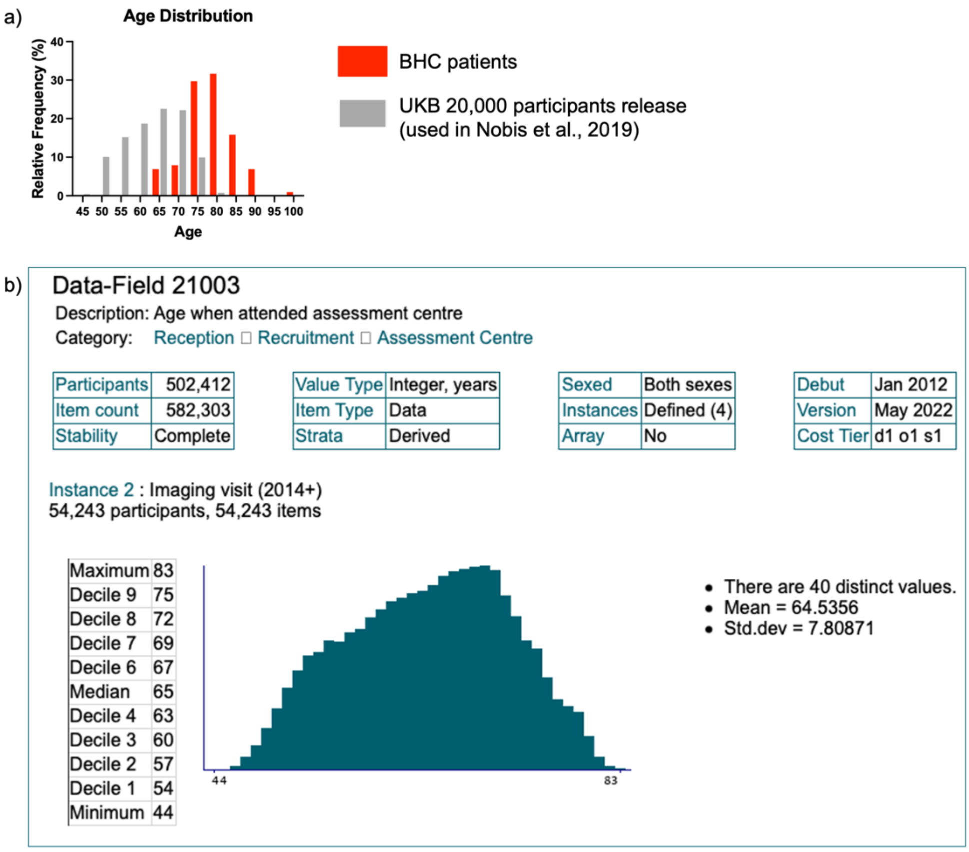


**Figure 3**. Age distribution in UK Biobank. a) Histogram of age distribution for 94 BHC patients (red) and 19,793 UKB participants from the 20,000 release (grey) used to build the nomograms (Nobis et al., 2019). b) Histogram of age distribution for UKB participants who underwent the imaging visit (release version May 2022, 54,243 participants - https://biobank.ndph.ox.ac.uk/showcase/field.cgi?id=21003). Also in the more recent release, the age range is different from BHC patients, highlighting need for expanded normative data.
